# The Impact of Obesity-Related Genetic Variants on NF-κB Signaling in Cardiovascular Disease: A Systematic Review

**DOI:** 10.1101/2024.12.11.24318855

**Authors:** Viqas Shafi, Nabeel Ahmad Khan, Ifrah Siddiqui

## Abstract

**Objective:** To investigate how genetic variations in obesity-related genes (FTO, MC4R, LEP, LEPR, PCSK1, PPARG, BDNF, SIM1, TBC1D1, ADRB3, UCP1, and SH2B1) impact the NF-κB signaling pathway, exploring mechanisms of NF-κB activation, downstream inflammatory responses, and their role in cardiovascular disease (CVD) development in the context of obesity.

**Background:** Obesity is a leading contributor to chronic diseases, including CVD, driven in part by chronic low-grade inflammation. The NF-κB signaling pathway, a central regulator of inflammatory responses, is implicated in the pathophysiology of obesity and CVD. Genetic variations in obesity-related genes may modulate NF-κB activation and its downstream effects, exacerbating inflammation and cardiovascular risks. Understanding these mechanisms can inform therapeutic strategies to mitigate inflammation and improve health outcomes in individuals with obesity.

**Methods:** A systematic review of the literature was conducted using PUBMED, MEDLINE, and Google Scholar, focusing on genetic variations in the obesity-related genes FTO, MC4R, LEP, LEPR, PCSK1, PPARG, BDNF, SIM1, TBC1D1, ADRB3, UCP1, and SH2B1. Studies examining the effects of these variations on NF-κB activation, inflammatory pathways, and CVD development were included. The search was performed with no date restrictions and followed PRISMA guidelines. Articles were screened for relevance, methodological rigor, and insights into the interplay between genetic factors, inflammation, and cardiovascular pathology. Data extraction focused on key findings linking gene variants to NF-κB signaling and their downstream effects.

**Results:** Genetic variations in FTO, MC4R, LEP, LEPR, and SH2B1 were found to disrupt insulin and leptin signaling, resulting in enhanced NF-κB activation and chronic inflammation. Variants in PPARG and UCP1 increased oxidative stress, further amplifying NF-κB signaling. These changes promoted endothelial dysfunction, atherosclerosis, and heightened CVD risk. Interactions between these genetic factors created a pro-inflammatory state, exacerbating cardiovascular complications in obese individuals.

**Conclusion:** This study underscores the critical role of genetic variations in obesity-related genes in modulating NF-κB signaling, driving chronic inflammation, and increasing CVD risk. Targeting these pathways may provide therapeutic opportunities to reduce inflammation and improve cardiovascular health in obese populations.

## Background

Obesity has emerged as one of the most pressing public health challenges of the 21st century, significantly increasing the risk of various chronic diseases, particularly cardiovascular disease (CVD). The World Health Organization estimates that over 1.9 billion adults were classified as overweight in 2021, with obesity affecting approximately 650 million individuals. The complex etiology of obesity encompasses genetic, environmental, and behavioral factors, yet the underlying mechanisms linking genetic predispositions to the pathophysiology of obesity-related CVD remain inadequately understood [1, 2].

Recent research has highlighted the critical role of inflammation in the development of obesity and its related complications. Chronic low-grade inflammation, driven by the activation of pro-inflammatory pathways such as NF-κB (nuclear factor kappa-light-chain-enhancer of activated B cells), has been implicated in the pathogenesis of CVD. The NF-κB signaling pathway mediates various cellular responses, including the regulation of immune responses, cell proliferation, and survival. Dysregulation of this pathway in the context of obesity can lead to persistent inflammation, contributing to endothelial dysfunction, atherosclerosis, and other cardiovascular pathologies [3, 4].

Given the growing prevalence of obesity and its associated health risks, it is essential to investigate the genetic variations in obesity-related genes—such as FTO, MC4R, LEP, LEPR, PCSK1, PPARG, BDNF, SIM1, TBC1D1, ADRB3, UCP1, and SH2B1—that influence NF-κB activation and inflammation. This study aims to elucidate the mechanisms by which these genetic factors contribute to the development of obesity and CVD. By enhancing our understanding of these relationships, we can inform targeted therapeutic strategies and develop personalized interventions to mitigate cardiovascular risks in obese populations. Ultimately, this research has the potential to improve health outcomes and reduce the burden of obesity-related diseases on individuals and healthcare systems [4, 5].

## Methods

### Aim of the Study

The aim of this study is to explore how genetic variations in obesity-related genes (FTO, MC4R, LEP, LEPR, PCSK1, PPARG, BDNF, SIM1, TBC1D1, ADRB3, UCP1, and SH2B1) influence the NF-κB signaling pathway. Specifically, the study investigates the mechanisms through which these genetic variations modulate NF-κB activation, its downstream inflammatory effects, and their contribution to cardiovascular disease (CVD) in the context of obesity.

### Research Question

How do genetic variations in key obesity-related genes impact the NF-κB signaling pathway and its role in inflammation-driven cardiovascular disease?

### Search Focus

A comprehensive literature search was conducted using PUBMED, MEDLINE, and Google Scholar, along with relevant open access and subscription-based journals. No date restrictions were applied to include the breadth of relevant research.

The search focused on the following key areas:

- **Obesity-Related Genes**: FTO, MC4R, LEP, LEPR, PCSK1, PPARG, BDNF, SIM1, TBC1D1, ADRB3, UCP1, SH2B1
- **Signaling Pathway**: NF-κB signaling and its downstream inflammatory mediators
- **Disease Context**: Cardiovascular disease and inflammation in obesity

The literature was screened and selected based on relevance to the study’s objectives. Literature search began in September 2021 and ended in May 2024. An in-depth investigation was conducted during this duration based on the parameters of the study as defined above. During revision, further literature was searched and referenced until November 2024. The literature search and all sections of the manuscript were checked multiple times during the months of revision (June 2024 – November 2024) to maintain the highest accuracy possible. This approach allowed for a comprehensive investigation on how genetic variations in key obesity-related genes impact the NF-κB signaling pathway and its role in inflammation-driven cardiovascular disease, adhering to the PRISMA guidelines for systematic reviews.

### Search Queries/Keywords

1. General Terms:

“Obesity-related genes” AND “NF-κB signaling” “Inflammation” AND “Cardiovascular disease” “Obesity” AND “Cardiovascular pathology”

2. Gene-Specific Searches: “FTO” AND “NF-κB”

“MC4R” AND “NF-κB signaling”

“LEP” OR “LEPR” AND “Inflammation” “PCSK1” AND “Obesity” AND “CVD”

“PPARG” AND “Cardiovascular disease” “BDNF” AND “Inflammatory pathways” “SIM1” AND “Obesity”

“TBC1D1” AND “NF-κB”

“ADRB3” AND “Inflammation” “UCP1” AND “NF-κB”

“SH2B1” AND “Obesity” AND “Inflammation”

3 Signaling and Disease-Specific Terms:

“NF-κB activation” AND “Cardiovascular disease” “Inflammation” AND “Obesity-related genes” “Inflammation” AND “Atherosclerosis”

“Chronic inflammation” AND “CVD” “Obesity-driven inflammation”

Boolean operators (AND, OR) were utilized to ensure comprehensive retrieval of relevant studies, capturing the intersections between genetic variations, NF-κB signaling, inflammation, and cardiovascular outcomes.

### Objectives of the Searches

- To explore how genetic variations in FTO, MC4R, LEP, LEPR, PCSK1, PPARG, BDNF, SIM1, TBC1D1, ADRB3, UCP1, and SH2B1 influence the NF-κB signaling pathway.
- To investigate the molecular mechanisms by which variations in obesity-related genes modulate NF-κB activation and downstream signaling.
- To examine how altered NF-κB signaling contributes to inflammation in the context of obesity, particularly focusing on chronic, low-grade inflammation as a driver of metabolic and cardiovascular complications.
- To evaluate the role of NF-κB-driven inflammatory pathways in the development and progression of cardiovascular disease in obese individuals.
- To synthesize data from various studies to provide a comprehensive understanding of the relationship between obesity-related genetic variations, NF-κB signaling, and cardiovascular disease.

#### Screening and Eligibility Criteria

- **Initial Screening**: Titles and abstracts were reviewed for direct relevance to the research question.
- **Full-Text Review**: Articles that passed initial screening were reviewed in full to ensure they provided detailed insights into the mechanisms of NF-κB activation influenced by genetic variations in obesity-related genes.

#### Inclusion Criteria

1. Studies exploring genetic variants in the targeted obesity-related genes.
2. Research focusing on the interaction between these genes and NF-κB signaling.
3. Articles linking altered NF-κB signaling to cardiovascular disease in obesity.
4. Peer-reviewed studies providing mechanistic insights into inflammation-driven CVD.

#### Exclusion Criteria

1. Studies lacking focus on the NF-κB pathway or its downstream effects.
2. Research with insufficient methodological rigor or unclear results.
3. Articles unrelated to obesity, genetic variations, or CVD.
4. Non-English publications and unpublished studies.

#### Data Extraction and Quality Assessment

Data extraction was focused on:

- Genetic variations and their functional effects on NF-κB signaling.
- Mechanistic insights into how these variations influence inflammatory markers.
- Evidence linking NF-κB dysregulation to cardiovascular outcomes.

To ensure reliability:

- **Quality Assessment**: Articles were assessed for methodological rigor, clear experimental designs, and peer-reviewed validation.
- **Bias Minimization**: Publication, selection, and reporting biases were addressed by diversifying sources and including studies with both positive and negative findings.

#### Rationale for Screening and Inclusion

- **FTO, MC4R, LEP, LEPR**: Linked to energy balance and inflammation, affecting NF-κB-mediated pathways.
- **PCSK1, PPARG**: Involved in metabolic regulation and chronic inflammation.
- **BDNF, SIM1**: Neural and metabolic roles influencing systemic inflammation.
- **TBC1D1, ADRB3**: Modulators of adipocyte function and inflammatory responses.
- **UCP1**: Thermogenic activity linked to anti-inflammatory pathways.
- **SH2B1**: Regulates insulin signaling, impacting inflammatory states in obesity.

Each gene was examined for its role in modulating NF-κB signaling, contributing to inflammation and cardiovascular pathology in obesity. This comprehensive analysis ensured a nuanced understanding of the genetic and molecular underpinnings of CVD in obesity.

#### Language and Publication Restrictions

Only English-language publications were included. No restrictions were placed on the date of publication to ensure a broad range of studies were considered. Unpublished studies, including conference abstracts, were excluded to prioritize peer-reviewed and validated research findings.

## Results

A total of 2104 articles were identified using database searching, and 2005 were recorded after duplicates removal. 1707 were excluded after screening of title/abstract, 168 were finally excluded, and 5 articles were excluded during data extraction. These exclusions were primarily due to factors such as non-conformity with the study focus, insufficient methodological rigor, or data that did not align with the research questions. Finally, 125 articles were included as references. PRISMA Flow Diagram is Fig 1.

**Fig 1.**
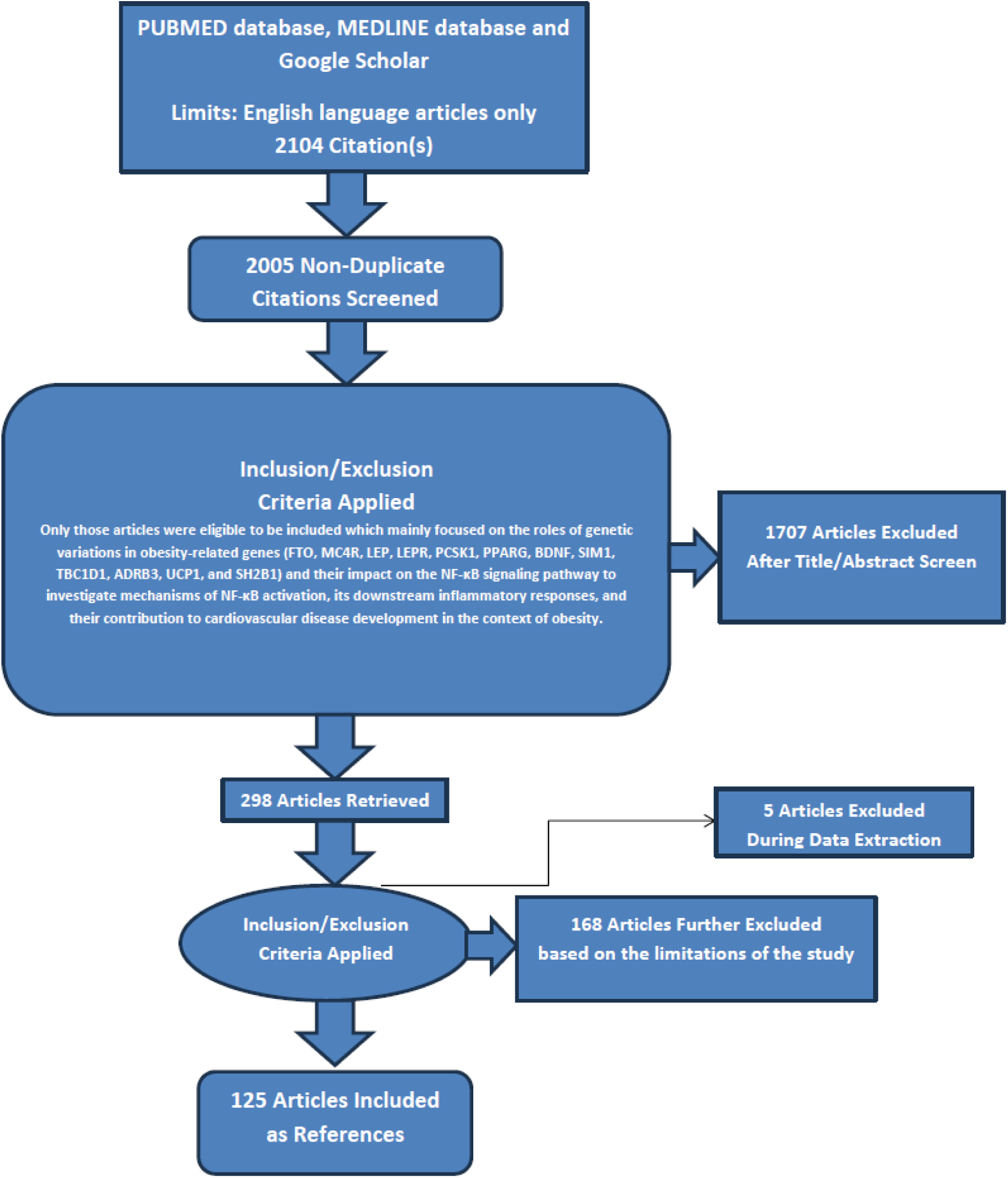
PRISMA FLOW DIAGRAM: This figure represents graphically the flow of citations in the study.

### Role of Obesity-Related Gene Variants in NF-κB Signaling and Cardiovascular Disease

#### 1. FTO (Fat Mass and Obesity-Associated) Gene

The FTO gene (Fat Mass and Obesity-Associated gene) is well-known for its role in obesity and body weight regulation. Recent studies have suggested that genetic variations in FTO can also impact the NF-κB signaling pathway, a key regulator of inflammation and immune response, which is often linked to cardiovascular disease (CVD) in the context of obesity [6].

##### FTO Variants and NF-κB Activation

Genetic variations in the fat mass and obesity-associated (FTO) gene have been closely examined for their roles in metabolic processes and inflammatory signaling pathways. Among these, the rs9939609 variant has garnered significant attention due to its association with altered expression and functionality of various metabolic genes. Evidence suggests that FTO variants can modulate NF-κB signaling by influencing adipocyte function, driving lipogenesis, and increasing the production of pro-inflammatory cytokines. These physiological changes can amplify NF-κB activation, a pathway critical for mediating inflammation [6, 7].

The role of FTO as an m6A RNA demethylase highlights its capacity to regulate gene expression through epigenetic mechanisms. By modifying mRNA stability and translation, FTO influences genes involved in the NF-κB pathway, including those encoding pro-inflammatory cytokines such as TNF-α and IL-6, as well as their receptors. Variants in FTO may enhance the stability and translational efficiency of these mRNAs, further contributing to the activation of NF-κB. This interaction underscores the gene’s potential impact on inflammatory responses and metabolic dysfunction [7, 8].

##### Mechanisms of NF-κB Activation by FTO Variants

FTO variations are linked to increased adiposity, leading to an enlarged adipose tissue mass that secretes pro-inflammatory adipokines such as TNF-α, IL-6, and MCP-1. These adipokines interact with surface receptors on adipocytes and immune cells, initiating signaling cascades that promote inflammation. This inflammatory environment further activates NF-κB, creating a feedback loop that sustains and amplifies the inflammatory response [8, 9].

Altered lipid metabolism is another pathway through which FTO influences NF-κB activation. Variants in FTO can contribute to dysregulated lipid handling, resulting in the production of reactive oxygen species (ROS). ROS serve as potent activators of NF-κB and exacerbate inflammatory processes, particularly in the cardiovascular system, highlighting the gene’s role in linking metabolic dysfunction to chronic inflammation [9, 10].

FTO variants also interact with other signaling pathways, such as those involved in insulin regulation. Dysregulated insulin signaling, often seen in obesity, can modulate NF-κB activity by elevating free fatty acid and glucose levels. These metabolic changes activate NF-κB through intermediates such as the IKK complex, establishing a molecular connection between metabolic dysregulation and inflammation [10, 11].

##### Downstream Effects of NF-κB Activation and Cardiovascular Disease (CVD)

NF-κB activation plays a critical role in the development of chronic inflammation and endothelial dysfunction. By upregulating the expression of adhesion molecules and cytokines, NF-κB facilitates the recruitment of immune cells to the vascular endothelium. This process contributes to the initiation and progression of atherosclerotic plaque formation, a central feature of cardiovascular disease [11, 12].

The interplay between FTO variants and NF-κB signaling can exacerbate lipid accumulation within vascular tissues. This pathological lipid deposition destabilizes atherosclerotic plaques, increasing the likelihood of adverse cardiovascular events such as myocardial infarction and stroke. The enhanced oxidative stress associated with this mechanism further amplifies tissue damage and inflammation, deepening the connection between metabolic dysfunction and vascular pathology.

Prolonged activation of NF-κB is also implicated in vascular remodeling and stiffening, processes commonly observed in obesity-associated hypertension. These structural and functional changes in the vasculature heighten cardiovascular risks, particularly in individuals with genetic susceptibility linked to FTO variants. The combined effects of inflammation, oxidative stress, and vascular dysfunction underscore the multidimensional role of NF-κB in mediating cardiovascular disease in this context [12, 13].

##### Implications for Cardiovascular Disease in the Context of Obesity

Altered NF-κB signaling associated with FTO variants drives a sustained cycle of inflammation, contributing to both systemic and vascular inflammatory processes. This chronic inflammatory state underpins key metabolic and vascular dysfunctions, including insulin resistance, dyslipidemia, and endothelial dysfunction. These conditions collectively represent significant risk factors for the development and progression of cardiovascular disease, particularly in the context of obesity [13, 14].

Insights into the role of FTO in modulating NF-κB signaling highlight potential avenues for therapeutic intervention. Strategies targeting the inflammatory cascade, such as the use of anti-inflammatory agents or NF- κB inhibitors, may reduce the impact of FTO-induced inflammation in obese individuals at high cardiovascular risk. Developing specific inhibitors of FTO presents an additional therapeutic approach, offering the potential to directly mitigate its pro-inflammatory and metabolic effects, thereby addressing a critical pathway in obesity-associated cardiovascular disease [14, 15].

#### 2. MC4R (Melanocortin-4 Receptor) Gene

Genetic variations in the MC4R (Melanocortin-4 Receptor) gene, primarily known for their impact on energy balance and appetite regulation, are also linked to altered NF-κB signaling, inflammation, and the development of cardiovascular disease (CVD) in the context of obesity [16].

##### MC4R Variants and NF-κB Activation

The melanocortin 4 receptor (MC4R) gene plays a critical role in regulating appetite and energy homeostasis within the hypothalamic melanocortin pathway. Variants of MC4R associated with obesity often lead to hyperphagia, characterized by excessive food intake, and decreased energy expenditure. Emerging evidence indicates that these genetic mutations extend their impact beyond metabolic regulation, influencing inflammatory pathways such as NF-κB signaling. The relationship appears to be mediated through indirect mechanisms linked to increased adiposity and associated inflammatory responses [16, 17].

Although MC4R itself does not directly activate NF-κB, the metabolic and inflammatory disturbances caused by obesity-related MC4R variants create conditions favorable for NF-κB pathway activation. The accumulation of adipose tissue in obese individuals promotes the secretion of pro-inflammatory cytokines such as tumor necrosis factor-alpha (TNF-α) and interleukin-6 (IL-6). These cytokines are potent activators of NF-κB, establishing a feedback loop that sustains chronic low-grade inflammation. This link between MC4R dysfunction, obesity, and inflammatory signaling highlights a complex interplay between genetic and environmental factors in the regulation of systemic inflammation [17, 18].

##### Mechanisms of NF-κB Activation by MC4R Variants

The deficiency of melanocortin 4 receptor (MC4R) function often results in excessive fat accumulation, a condition tightly linked to increased adipose tissue inflammation. Excess fat promotes the release of pro-inflammatory cytokines, including tumor necrosis factor-alpha (TNF-α) and interleukin-6 (IL-6). These cytokines activate NF-κB through receptor-mediated pathways involving Toll-like receptors (TLRs) and the TNF receptor, triggering a systemic inflammatory response. This inflammation contributes to the broader metabolic and immune dysregulation observed in individuals with MC4R variants [18, 19].

In addition to adipose tissue inflammation, MC4R dysfunction can influence sympathetic nervous system (SNS) activity. Normally, MC4R regulates SNS signaling to maintain energy balance and cardiovascular homeostasis. Variants that impair MC4R function disrupt this regulation, potentially causing excessive catecholamine release. Elevated levels of stress hormones such as epinephrine and norepinephrine can indirectly activate NF-κB in cardiovascular tissues, fostering inflammation and contributing to endothelial dysfunction [19, 20].

MC4R variants are also associated with cellular stress responses, particularly endoplasmic reticulum (ER) stress, which is prevalent in adipose tissue and the liver. ER stress activates the unfolded protein response (UPR), a mechanism that can stimulate NF-κB signaling. The resulting inflammatory cascade exacerbates tissue dysfunction and metabolic disturbances, establishing a link between MC4R-related genetic defects and chronic inflammation. These interconnected mechanisms underscore the multidimensional impact of MC4R variants on inflammatory pathways and systemic health [20, 21].

##### Downstream Effects of NF-κB Activation and Cardiovascular Disease (CVD)

Chronic inflammation induced by MC4R variants leads to NF-κB activation, which plays a critical role in the development of atherosclerosis. The inflammatory environment enhances the production of cytokines and chemokines, which in turn attract immune cells to the endothelial layer of blood vessels. This process accelerates the formation of lipid-laden plaques, contributing to the thickening and stiffening of arterial walls, a hallmark of atherosclerosis. Over time, this can lead to reduced blood flow and increased risk of cardiovascular events [21,22].

NF-κB-driven inflammation also impacts endothelial cell function, a critical element in maintaining vascular health. Endothelial dysfunction, a consequence of persistent inflammation, is a well-established precursor to hypertension and other cardiovascular diseases (CVD). This dysfunction reduces the bioavailability of nitric oxide, a key molecule responsible for vascular dilation, while simultaneously promoting increased vascular stiffness. The resulting impairment in vascular tone and elasticity elevates the risk of hypertension, further exacerbating the development of CVD [22, 23].

Obesity related to MC4R dysfunction is frequently accompanied by insulin resistance, a condition that also stimulates NF-κB activation. This occurs through mechanisms involving advanced glycation end-products (AGEs) and reactive oxygen species (ROS), both of which enhance inflammatory signaling pathways. Additionally, dyslipidemia, characterized by elevated levels of cholesterol and triglycerides, is commonly observed in individuals with MC4R variants. This lipid imbalance is closely linked to NF-κB activation and represents a significant risk factor for the development of CVD, further highlighting the complex interplay between obesity, inflammation, and cardiovascular health [23, 24].

##### Implications for Cardiovascular Disease in the Context of Obesity

The increased susceptibility to cardiovascular disease (CVD) in individuals with MC4R variants, especially when associated with obesity, is largely mediated through NF-κB-driven inflammation. This chronic inflammatory response plays a crucial role in the pathogenesis of atherosclerosis, hypertension, and other cardiovascular conditions, which significantly elevates the risk of adverse cardiovascular events in this population [24, 25].

Considering the role of MC4R in modulating NF-κB and its impact on cardiovascular health, there is potential for targeted therapeutic strategies. MC4R agonists, which enhance the function of the melanocortin 4 receptor, could offer a way to mitigate obesity-related inflammation and consequently reduce the risk of CVD. Additionally, anti-inflammatory agents that specifically target the NF-κB pathway may provide therapeutic benefits by controlling systemic inflammation and reducing the incidence of CVD in individuals with MC4R-related obesity.

These approaches could help break the cycle of chronic inflammation and cardiovascular pathology associated with MC4R dysfunction, offering a new avenue for managing obesity and its cardiovascular complications [25].

#### 3. LEP (Leptin) Gene

Genetic variations in the LEP (Leptin) gene, which encodes the hormone leptin, play a crucial role in regulating energy balance and fat storage. These variations can also impact the NF-κB signaling pathway, a key regulator of inflammation, which contributes to cardiovascular disease (CVD) in the context of obesity [26].

##### LEP Variants and NF-κB Activation

Genetic variations in the LEP gene, which encodes the hormone leptin, can lead to either leptin deficiency or altered leptin signaling, both of which influence the NF-κB pathway. Leptin plays a significant role in regulating energy balance and inflammation by activating NF-κB through its receptors present on a variety of cells, including immune cells and endothelial cells. This activation triggers inflammatory responses that are central to the development of metabolic and cardiovascular diseases [26, 27].

In the context of obesity, LEP gene variants frequently contribute to a phenomenon known as leptin resistance. In this condition, despite elevated levels of leptin, the signaling pathways mediated by leptin are impaired, leading to a disruption in its normal regulatory functions. Leptin resistance fosters chronic low-grade inflammation, which is a key driver of NF-κB activation. This persistent inflammatory state primarily affects adipose tissue and the cardiovascular system, further exacerbating the development of obesity-related complications, including atherosclerosis, hypertension, and other forms of cardiovascular disease [27, 28].

##### Mechanisms of NF-κB Activation by LEP Variants

Leptin, through its receptor (LEPR), directly stimulates NF-κB activation by initiating signaling cascades that involve the Janus kinase (JAK) and signal transducer and activator of transcription (STAT) pathways. These pathways can intersect with NF-κB signaling, leading to the activation of pro-inflammatory genes. In individuals with LEP gene variations that result in elevated leptin levels, this signaling cascade is amplified, promoting the expression of pro-inflammatory cytokines such as tumor necrosis factor-alpha (TNF-α) and interleukin-6 (IL-6). These cytokines contribute to systemic inflammation, further exacerbating the risk of metabolic and cardiovascular disorders [28, 29].

Leptin also plays a key role in attracting and activating macrophages within adipose tissue. Once activated, macrophages release additional inflammatory cytokines that further stimulate NF-κB, establishing a self-reinforcing inflammatory loop. This pro-inflammatory environment in adipose tissue amplifies local inflammation and enhances NF-κB signaling within both adipose and vascular tissues. As a result, the chronic inflammation in these tissues can drive the progression of obesity-related complications, such as atherosclerosis and hypertension [29, 30].

In addition to its effects on immune cells, elevated leptin also increases oxidative stress by promoting the production of reactive oxygen species (ROS). ROS can activate the IκB kinase (IKK) complex, a critical regulator of NF-κB activation. The subsequent NF-κB activation contributes to inflammation and endothelial dysfunction, particularly in the cardiovascular system. This cascade of oxidative stress and inflammation underscores the potential for leptin-related signaling to accelerate the development of cardiovascular diseases in individuals with LEP gene variants [30, 31].

##### Downstream Effects of NF-κB Activation and Cardiovascular Disease (CVD)

Chronic activation of NF-κB in response to leptin-induced inflammation plays a crucial role in the development of atherosclerosis. NF-κB promotes the expression of adhesion molecules and chemokines that recruit immune cells to the endothelial layer of blood vessels. These immune cells contribute to the formation of atherosclerotic plaques, thickening and stiffening arterial walls, which can obstruct blood flow and increase the risk of cardiovascular events [31, 32].

Leptin’s effects on endothelial function are also significant, as it impairs the ability of endothelial cells to maintain vascular health. Through NF-κB-mediated inflammatory pathways, leptin reduces the bioavailability of nitric oxide, a molecule essential for vasodilation and proper blood flow. Impaired endothelial function is a key factor in the development of hypertension and other cardiovascular disorders, as it disrupts the balance between vasoconstriction and vasodilation, leading to elevated blood pressure [32, 33].

Furthermore, NF-κB activation in the vascular endothelium due to elevated leptin levels promotes vascular remodeling and fibrosis. These processes contribute to arterial stiffness, which in turn exacerbates hypertension. As arterial walls become less elastic, the heart must work harder to pump blood, further raising blood pressure. The combination of vascular remodeling, fibrosis, and NF-κB-induced inflammation creates a cycle that intensifies cardiovascular risk, underscoring the impact of leptin-related signaling in the pathophysiology of cardiovascular disease [33, 34].

##### Implications for Cardiovascular Disease in the Context of Obesity

Leptin-induced activation of NF-κB plays a central role in the chronic inflammation observed in obesity, which is closely associated with cardiovascular disease (CVD). The interaction between leptin, NF-κB, and inflammation highlights the critical role of these pathways in driving cardiovascular risk in obese individuals. Chronic low-grade inflammation in adipose tissue, coupled with the systemic effects of elevated leptin, exacerbates the development of atherosclerosis, endothelial dysfunction, and hypertension. Targeting this inflammatory axis could significantly reduce the cardiovascular burden in individuals with obesity [34, 35].

Therapeutic strategies aimed at reducing NF-κB activation or inhibiting leptin signaling offer promising avenues for managing obesity-related cardiovascular risk. NF-κB inhibitors or anti-inflammatory agents that specifically target leptin’s effects on inflammation may help mitigate the progression of CVD in this population. Additionally, lifestyle interventions that lower leptin levels, such as weight loss and regular exercise, could also reduce NF-κB- driven inflammation and improve cardiovascular health. These approaches emphasize the importance of addressing the underlying inflammatory mechanisms to improve long-term cardiovascular outcomes in individuals with obesity [35].

#### 4. LEPR (Leptin Receptor) Gene

Genetic variations in the LEPR (Leptin Receptor) gene, which encodes the receptor for the hormone leptin, are closely associated with obesity and related metabolic dysfunctions. These variations can influence NF-κB (nuclear factor kappa-light-chain-enhancer of activated B cells) signaling, which plays a central role in inflammation and cardiovascular disease (CVD) development [36].

##### LEPR Variants and NF-κB Activation

Genetic variants in the LEPR gene, which encodes the leptin receptor, can result in leptin resistance, a condition where the body fails to respond appropriately to leptin despite elevated hormone levels. This resistance impairs leptin’s ability to regulate energy balance, and more critically, it promotes a chronic state of low-grade inflammation. In this inflammatory environment, NF-κB signaling becomes activated, further exacerbating metabolic dysregulation and contributing to the development of obesity-related complications [36, 37].

LEPR variants are often linked to increased adiposity, which leads to enhanced leptin production. However, the altered function of the leptin receptor in these variants can intensify leptin’s pro-inflammatory actions. The dysfunction of LEPR not only disrupts normal metabolic regulation but also amplifies NF-κB activation in a variety of tissues, particularly in adipose and vascular tissues. This heightened NF-κB signaling perpetuates a cycle of inflammation, contributing to the pathogenesis of obesity-related diseases such as atherosclerosis and hypertension. The interplay between LEPR variants, leptin resistance, and NF-κB activation underscores the critical role of leptin receptor signaling in the progression of chronic inflammation and cardiovascular disease [37,38].

##### Mechanisms of NF-κB Activation by LEPR Variants

Leptin normally binds to the leptin receptor (LEPR), triggering signaling pathways that involve Janus kinase 2 (JAK2) and signal transducer and activator of transcription 3 (STAT3). These pathways can intersect with NF-κB signaling, allowing leptin to regulate both energy balance and inflammatory responses. However, variants in LEPR that lead to leptin resistance result in prolonged and dysregulated leptin signaling. This impaired signaling contributes to heightened activation of NF-κB, as it stimulates the release of inflammatory cytokines such as tumor necrosis factor-alpha (TNF-α) and interleukin-6 (IL-6), which further amplify inflammatory pathways [38,39].

Leptin resistance, often associated with LEPR mutations, also leads to increased macrophage infiltration in adipose tissue. These macrophages produce additional pro-inflammatory cytokines that activate NF-κB, establishing a feedback loop of inflammation. This cycle perpetuates inflammation not only in adipose tissue but also in the vasculature, creating an environment that accelerates metabolic dysfunction and the development of cardiovascular diseases [39, 40].

Additionally, leptin signaling through LEPR can directly activate NF-κB in endothelial cells, particularly when leptin levels are elevated. This activation induces the expression of adhesion molecules and other inflammatory mediators that contribute to endothelial dysfunction. LEPR variants that disrupt normal receptor function exacerbate this process, leading to further endothelial dysfunction and inflammation, which are key factors in the progression of cardiovascular disease (CVD). The combined effect of altered leptin receptor signaling, increased inflammation, and endothelial dysfunction underscores the critical role of LEPR variants in the pathogenesis of obesity-related CVD [40, 41].

##### Downstream Effects of NF-κB Activation and Cardiovascular Disease (CVD)

Chronic activation of NF-κB, resulting from dysregulated leptin receptor (LEPR) signaling, plays a significant role in promoting systemic inflammation, which is closely linked to the development of atherosclerosis. This prolonged inflammation drives the release of pro-inflammatory cytokines, leading to the recruitment of immune cells to the endothelial surface of blood vessels. These immune cells contribute to plaque formation, further enhancing vascular inflammation. Over time, this inflammatory process contributes to the thickening and stiffening of arterial walls, a hallmark of atherosclerosis, increasing the risk of cardiovascular events [41, 42].

In addition to inflammation, dysregulated leptin signaling through LEPR variants also leads to increased production of reactive oxygen species (ROS), resulting in oxidative stress. ROS can activate the NF-κB pathway, which in turn contributes to vascular remodeling and arterial stiffness. These changes in vascular structure are directly linked to the development of hypertension, a major risk factor for cardiovascular disease (CVD). The combination of increased oxidative stress, vascular remodeling, and NF-κB activation creates a vicious cycle that accelerates CVD progression [42, 43].

LEPR variants are often associated with obesity-related hypertension, in part due to the inflammation mediated by NF-κB. This chronic inflammatory state also plays a role in the development of insulin resistance and dyslipidemia, conditions that further compound the risk of cardiovascular complications. As a result, individuals with dysregulated leptin receptor signaling face an increased likelihood of metabolic syndrome, which significantly amplifies their risk of developing CVD. The interconnected nature of these pathways underscores the central role of leptin receptor variants in driving obesity-related cardiovascular and metabolic disorders [43, 44].

##### Implications for Cardiovascular Disease in the Context of Obesity

The interaction between LEPR variants, leptin resistance, and NF-κB activation represents a critical pathway through which obesity promotes cardiovascular disease (CVD). Dysregulated leptin signaling leads to chronic low-grade inflammation, which is a central factor in the development of atherosclerosis, endothelial dysfunction, and metabolic syndrome—key precursors to CVD. The persistent inflammatory state driven by NF-κB exacerbates these conditions, contributing to the accelerated progression of cardiovascular complications in obese individuals with leptin resistance [44, 45].

Given the central role of NF-κB in this inflammatory cascade, targeting its signaling pathway and the inflammatory mediators it activates offers a promising therapeutic strategy. Anti-inflammatory drugs or specific NF-κB inhibitors could help mitigate the inflammatory burden in individuals with LEPR-related leptin resistance. In addition to pharmacological interventions, lifestyle changes that reduce adiposity and improve leptin sensitivity, such as weight loss and increased physical activity, may further alleviate inflammation and improve cardiovascular outcomes. These approaches highlight the potential for both pharmacological and lifestyle-based therapies to reduce the cardiovascular risks associated with obesity-related leptin resistance [45].

#### 5. PCSK1 (Proprotein Convertase Subtilisin/Kexin Type 1) Gene

The PCSK1 (Proprotein Convertase Subtilisin/Kexin Type 1) gene encodes an enzyme essential for processing various precursor proteins, including those involved in metabolic regulation. Genetic variations in PCSK1 have been associated with obesity and related metabolic disturbances. These variations can indirectly influence NF-κB (nuclear factor kappa-light-chain-enhancer of activated B cells) signaling, which plays a significant role in inflammation and cardiovascular disease (CVD) [46].

##### PCSK1 Variants and NF-κB Activation

The proprotein convertase subtilisin/kexin type 1 (PCSK1) plays a critical role in the activation of several pro-inflammatory cytokines and hormones. These include proinsulin, proglucagon, and peptides derived from proopiomelanocortin (POMC). Variants in PCSK1 can interfere with these processing events, resulting in an imbalance of metabolic and inflammatory mediators. This imbalance has the potential to indirectly enhance nuclear factor-kappa B (NF-κB) signaling, a pathway central to inflammation and immune responses [46, 47].

PCSK1 variations have also been implicated in metabolic dysregulation, with a strong association with early-onset obesity and hyperphagia. Impaired processing of key metabolic hormones, such as insulin and glucagon-like peptides, contributes to this phenotype. These metabolic disturbances often lead to chronic low-grade inflammation, a condition that is closely linked to the activation of NF-κB, particularly in adipose and vascular tissues. This suggests that PCSK1 variants may serve as a molecular bridge between metabolic dysfunction and inflammatory signaling, highlighting their relevance in the pathophysiology of obesity and related conditions [47,48].

##### Mechanisms of NF-κB Activation by PCSK1 Variants

Mutations in PCSK1 disrupt the processing of proopiomelanocortin (POMC)-derived peptides, including α- melanocyte-stimulating hormone (α-MSH), a key regulator of appetite and energy balance. Deficiencies in these peptides contribute to obesity and adipose tissue inflammation, conditions that foster the release of pro-inflammatory cytokines such as interleukin-6 (IL-6) and tumor necrosis factor-alpha (TNF-α). These cytokines can activate the NF-κB signaling pathway through engagement with receptors such as TNF receptors (TNFR) and Toll-like receptors (TLR), amplifying inflammatory responses [48, 49].

PCSK1 variants are also associated with increased lipid accumulation in adipose tissue, which can attract macrophage infiltration. These infiltrating macrophages secrete additional pro-inflammatory cytokines, further driving NF-κB activation and creating a self-sustaining inflammatory loop within adipose and cardiovascular tissues. This feedback mechanism exacerbates metabolic and inflammatory imbalances, potentially contributing to the progression of obesity-related comorbidities [49, 50].

Additionally, PCSK1 mutations may induce endoplasmic reticulum (ER) stress due to the accumulation of misprocessed proteins. This stress activates the unfolded protein response (UPR) pathway, a mechanism known to stimulate NF-κB. The resulting inflammation, coupled with oxidative stress, may intensify systemic inflammation and heighten cardiovascular risk, particularly in the context of obesity. These mechanisms underscore the multidimensional role of PCSK1 in linking metabolic dysregulation to inflammatory pathways [50,51].

##### Downstream Effects of NF-κB Activation and Cardiovascular Disease

Inflammation mediated by NF-κB activation plays a critical role in the development of atherosclerosis, particularly in the context of PCSK1-related dysfunction. This activation promotes the expression of adhesion molecules and chemokines, which facilitate the recruitment of immune cells to the vascular endothelium. The accumulation of these cells accelerates the formation and progression of atherosclerotic plaques, a critical precursor to cardiovascular events such as myocardial infarction and stroke [51, 52].

Chronic NF-κB-driven inflammation also contributes to endothelial dysfunction by diminishing nitric oxide bioavailability, an essential factor for maintaining vascular health and promoting vasodilation. Reduced nitric oxide levels impair endothelial cell function, promoting vasoconstriction and increasing the risk of hypertension, a well-established contributor to cardiovascular disease [52, 53].

Metabolic dysregulation associated with PCSK1 mutations exacerbates these processes through its links to insulin resistance and dyslipidemia. Insulin resistance elevates circulating glucose and free fatty acid levels, which further stimulate NF-κB activity, amplifying inflammatory responses. Concurrently, dyslipidemia fosters the deposition of lipids within arterial walls, directly contributing to atherogenesis. Together, these interconnected mechanisms underscore the role of PCSK1 variants and NF-κB activation in driving cardiovascular disease risk [53,54].

##### Implications for Cardiovascular Disease in the Context of Obesity

The interplay between PCSK1 variants, metabolic dysregulation, and NF-κB activation highlights the critical role of chronic inflammation in the pathogenesis of obesity-related cardiovascular disease. NF-κB-driven inflammation exacerbates key processes such as atherosclerosis, endothelial dysfunction, and the broader manifestations of metabolic syndrome, each of which contributes to cardiovascular pathology. This underscores the importance of addressing inflammation as a central mechanism linking obesity to cardiovascular disease [54, 55].

Targeting these pathways presents potential therapeutic opportunities. Interventions aimed at reducing NF-κB activation or mitigating its downstream inflammatory effects could be particularly beneficial for individuals with PCSK1-related obesity. Anti-inflammatory agents, either directly targeting NF-κB or modulating its effector pathways, may help alleviate the inflammatory burden associated with metabolic and cardiovascular dysregulation. Enhancing the processing of metabolic hormones affected by PCSK1 mutations represents another promising strategy. Complementary lifestyle interventions focused on reducing adiposity and improving overall metabolic health could further attenuate NF-κB-mediated cardiovascular risk, offering a holistic approach to managing obesity-related cardiovascular disease [55].

#### 6. PPARG (Peroxisome Proliferator-Activated Receptor Gamma) Gene

Genetic variations in the PPARG (Peroxisome Proliferator-Activated Receptor Gamma) gene, which plays a critical role in adipogenesis, glucose metabolism, and lipid homeostasis, are implicated in obesity and metabolic disorders. These variations can influence NF-κB (nuclear factor kappa-light-chain-enhancer of activated B cells) signaling, a key regulator of inflammation and cardiovascular disease (CVD) [56].

##### PPARG Variants and NF-κB Activation

The peroxisome proliferator-activated receptor gamma (PPARG) plays a crucial role in regulating inflammation by suppressing the activity of nuclear factor-kappa B (NF-κB). This anti-inflammatory function helps maintain homeostasis in various tissues, including adipose and vascular cells. Variants in the PPARG gene can impair its regulatory capacity, reducing its ability to inhibit NF-κB activity and promoting a shift toward a pro-inflammatory state. This heightened inflammatory signaling can exacerbate systemic metabolic and vascular disturbances [56,57].

PPARG variants are strongly associated with metabolic conditions such as obesity, insulin resistance, and type 2 diabetes. These disorders are characterized by chronic low-grade inflammation, which is mediated in part by increased NF-κB activation in adipose and vascular tissues. Insulin resistance further amplifies this cycle by elevating levels of circulating free fatty acids and glucose, both of which are known activators of NF-κB. Together, these mechanisms link PPARG dysfunction to the interplay between metabolic dysregulation and inflammation, contributing to the pathogenesis of obesity-related and vascular diseases [57, 58].

##### Mechanisms of NF-κB Activation by PPARG Variants

PPARG serves as a critical modulator of the NF-κB signaling pathway, largely through its role in promoting the expression of IκBα, a key inhibitor of NF-κB. This regulatory function suppresses NF-κB activation and limits the production of pro-inflammatory cytokines such as tumor necrosis factor-alpha (TNF-α) and interleukin-6 (IL-6). Loss-of-function variants in PPARG compromise this inhibitory mechanism, leading to unchecked NF-κB activation and increased pro-inflammatory signaling [58, 59].

In adipose tissue, PPARG variants disrupt normal adipocyte differentiation and lipid storage, resulting in adipose tissue expansion and inflammation. This dysfunctional state promotes the secretion of inflammatory cytokines, further activating NF-κB and perpetuating a cycle of inflammation. The resulting chronic low-grade inflammation exacerbates obesity-related complications, including insulin resistance and metabolic syndrome [59, 60].

PPARG also plays a vital role in endothelial cell function, where it helps regulate oxidative stress and inflammatory responses. Variants that impair PPARG activity contribute to endothelial dysfunction, increasing oxidative stress and activating NF-κB in vascular tissues. This promotes vascular inflammation, which is a precursor to the development of atherosclerosis and other cardiovascular diseases. These mechanisms emphasize the significance of PPARG variants in linking metabolic dysfunction to inflammation and vascular pathology [60, 61].

##### Downstream Effects of NF-κB Activation and Cardiovascular Disease

Dysfunction in PPARG contributes to NF-κB activation in vascular tissues, driving the expression of adhesion molecules and inflammatory mediators. These factors facilitate the recruitment of immune cells to the vascular endothelium, initiating and sustaining vascular inflammation. Over time, this process accelerates the development of atherosclerotic plaques, which are major contributors to cardiovascular disease (CVD), including heart attacks and strokes [62, 63].

PPARG variants are closely associated with insulin resistance, a condition that heightens the risk of hypertension and exacerbates NF-κB activation. Insulin resistance increases oxidative stress and promotes the production of inflammatory cytokines, further fueling NF-κB signaling. The combination of heightened inflammation and elevated blood pressure places additional strain on the cardiovascular system, increasing the likelihood of vascular damage and CVD [63, 64].

Impaired PPARG function also disrupts lipid metabolism, resulting in dyslipidemia characterized by elevated levels of circulating lipids. These lipids, when oxidized, act as potent activators of NF-κB in endothelial cells. This activation enhances inflammatory signaling and promotes lipid deposition within arterial walls, driving the progression of atherosclerotic plaques. Together, these mechanisms underscore the interconnected roles of PPARG dysfunction, NF-κB activation, and metabolic disturbances in the pathogenesis of cardiovascular disease [64, 65].

##### Implications for Cardiovascular Disease in the Context of Obesity

The interplay between PPARG variants, metabolic dysregulation, and NF-κB activation underscores the critical role of inflammation in the development of obesity-related cardiovascular disease (CVD). Chronic NF-κB-driven inflammation is a key driver of atherosclerosis, endothelial dysfunction, and metabolic syndrome, all of which contribute to cardiovascular pathology. The heightened inflammatory state associated with PPARG dysfunction links metabolic imbalances directly to vascular and cardiac complications, emphasizing the importance of addressing these interconnected mechanisms in CVD management [64].

Therapeutic strategies aimed at enhancing PPARG activity hold promise for individuals with PPARG-related metabolic dysfunction. PPARG agonists, such as thiazolidinediones (TZDs), have shown potential in restoring PPARG function, improving insulin sensitivity, and reducing NF-κB activation. These interventions could help mitigate inflammation and metabolic disturbances, addressing a root cause of CVD. Additionally, anti-inflammatory agents targeting NF-κB signaling and its downstream pathways offer another avenue to reduce inflammation and associated vascular damage. Lifestyle modifications, including weight loss, improved diet, and increased physical activity, complement pharmacological approaches by alleviating metabolic stress and reducing inflammation, further lowering cardiovascular risk [65].

#### 7. BDNF (Brain-Derived Neurotrophic Factor) Gene

Genetic variations in the BDNF (Brain-Derived Neurotrophic Factor) gene, which encodes a protein involved in neuronal growth, differentiation, and energy homeostasis, have been associated with obesity, metabolic disturbances, and increased risk for cardiovascular disease (CVD). These variations can influence NF-κB (nuclear factor kappa-light-chain-enhancer of activated B cells) signaling, a pathway known for its role in inflammation and cardiovascular pathology [66].

##### BDNF Variants and NF-κB Activation

Brain-derived neurotrophic factor (BDNF) is a key regulator of appetite, energy expenditure, and neuronal plasticity. Variants in the BDNF gene, such as the Val66Met polymorphism, can lead to reduced expression or altered activity of BDNF. This deficiency is associated with an increased propensity for obesity and a heightened pro-inflammatory state, conditions that are strongly linked to the activation of nuclear factor-kappa B (NF-κB) signaling. Reduced BDNF levels disrupt normal energy homeostasis, contributing to metabolic dysregulation and systemic inflammation [66, 67].

BDNF deficiency, often resulting from genetic variations, contributes to obesity by enhancing appetite and decreasing energy expenditure. This excess adiposity triggers a cascade of inflammatory processes, particularly within adipose tissue, where the secretion of cytokines such as TNF-α and IL-6 is upregulated. These cytokines activate NF-κB in adipose and vascular tissues, creating a feedback loop that sustains chronic inflammation. This interplay between BDNF variants, obesity, and NF-κB activation highlights a critical mechanism linking metabolic dysfunction to inflammation and associated health risks [67, 68].

##### Mechanisms of NF-κB Activation by BDNF Variants

Reduced BDNF activity has a direct impact on energy balance, contributing to increased food intake and decreased energy expenditure, both of which promote fat accumulation and obesity. The resulting excess adiposity triggers the release of pro-inflammatory cytokines, such as TNF-α and IL-6. These cytokines activate NF- κB, leading to a pro-inflammatory environment that exacerbates metabolic dysfunction and accelerates the progression of obesity-related complications. The dysregulated energy balance thus creates a feedback loop where NF-κB signaling further amplifies the inflammatory response [68, 69].

BDNF variants also influence neuroimmune communication by modulating neuronal pathways that regulate immune responses. When BDNF levels are reduced, inflammation in the central nervous system (CNS) can spill over into peripheral systems, elevating cytokine production and further activating NF-κB signaling. This cross-talk between the nervous and immune systems enhances systemic inflammation, perpetuating the inflammatory milieu associated with obesity and metabolic disorders [69, 70].

Moreover, BDNF plays a critical role in managing oxidative stress within both neuronal and non-neuronal tissues. Genetic variants that lead to reduced BDNF levels increase oxidative stress by elevating reactive oxygen species (ROS). These ROS directly activate NF-κB signaling pathways, contributing to cellular damage and inflammation, particularly in the cardiovascular system. The combined effects of oxidative stress and NF-κB activation underscore the potential cardiovascular risks associated with BDNF deficiency [70, 71].

##### Downstream Effects of NF-κB Activation and Cardiovascular Disease (CVD)

Chronic NF-κB activation resulting from BDNF deficiency contributes to endothelial dysfunction by promoting the expression of adhesion molecules and inflammatory mediators in vascular endothelial cells. These molecules facilitate the recruitment of immune cells to the endothelium, initiating the formation of plaques and the progression of atherosclerosis. This process is central to the development of cardiovascular diseases, as the inflammatory environment in the vascular walls accelerates plaque buildup, narrowing arteries and increasing the risk of heart attacks and strokes [71, 72].

BDNF deficiency, through its connection to obesity, also plays a significant role in the development of hypertension. The NF-κB-mediated inflammation and resulting endothelial dysfunction increase vascular resistance, contributing to elevated blood pressure. Chronic inflammation in the vascular endothelium impairs normal blood vessel function, leading to persistent hypertension, which is a major risk factor for cardiovascular events such as heart failure and stroke [72, 73].

In addition, BDNF variants associated with obesity and metabolic dysregulation promote insulin resistance, partly through enhanced NF-κB activation. Insulin resistance exacerbates systemic inflammation and increases circulating glucose and free fatty acids, further stimulating NF-κB signaling. This creates a vicious cycle of metabolic dysfunction and inflammation, which not only worsens insulin resistance but also elevates the risk of developing cardiovascular diseases, reinforcing the link between metabolic and vascular health [73, 74].

##### Implications for Cardiovascular Disease in the Context of Obesity

The interaction between BDNF variants, metabolic dysregulation, and NF-κB signaling highlights the critical role of inflammation in the development of obesity-related cardiovascular disease (CVD). Chronic NF-κB-driven inflammation exacerbates key processes such as atherosclerosis, endothelial dysfunction, and the broader spectrum of metabolic syndrome, all of which contribute to the progression of cardiovascular pathology. The inflammatory environment fueled by BDNF deficiency accelerates vascular damage, increasing the risk of heart disease, stroke, and other cardiovascular events [74, 75].

Therapeutic strategies aimed at enhancing BDNF activity or reducing NF-κB activation may provide significant benefits for individuals with BDNF-related obesity and associated cardiovascular risk. One promising approach is lifestyle interventions, particularly exercise, which has been shown to increase BDNF levels and help reduce inflammation. In addition to these behavioral changes, pharmacological agents targeting NF-κB signaling and improving metabolic health may further mitigate inflammation and its associated cardiovascular risks. Together, these interventions could offer a comprehensive approach to addressing obesity-related cardiovascular disease [75].

#### 8. SIM1 (Single-Minded Homolog 1) Gene

Genetic variations in the SIM1 (Single-Minded Homolog 1) gene, a transcription factor primarily involved in the development of the hypothalamus and regulation of appetite, have been associated with obesity and metabolic disturbances. Variations in this gene can indirectly influence the NF-κB (nuclear factor kappa-light-chain-enhancer of activated B cells) signaling pathway, which is central to inflammation and cardiovascular disease (CVD) [76].

##### SIM1 Variants and NF-κB Activation

SIM1 (single-minded 1) plays a critical role in the normal development and function of hypothalamic regions involved in regulating hunger and satiety. Variants in SIM1 can disrupt this regulatory function, leading to hyperphagia (excessive eating) and obesity. These conditions are often associated with chronic low-grade inflammation, a key driver of enhanced NF-κB activation. The dysregulation of energy homeostasis due to SIM1 mutations creates a feedback loop where increased food intake and obesity promote inflammation, further stimulating NF-κB signaling [76, 77].

The obesity associated with SIM1 dysfunction also leads to adipose tissue inflammation. This inflammatory response triggers the release of pro-inflammatory cytokines such as TNF-α and IL-6, which activate NF-κB signaling pathways. Persistent activation of NF-κB exacerbates systemic inflammation, contributing to metabolic dysregulation and increasing the risk of cardiovascular complications. This link between SIM1 variants, obesity, and chronic inflammation underscores the importance of understanding how hypothalamic dysfunction can influence systemic health, particularly in relation to cardiovascular disease [77, 78].

##### Mechanisms of NF-κB Activation by SIM1 Variants

Variants in SIM1 can lead to altered hypothalamic signaling, resulting in overeating and obesity. The increased adiposity that accompanies SIM1-related obesity promotes inflammatory responses, particularly within adipose tissue. These inflammatory responses involve the release of pro-inflammatory cytokines such as IL-6 and TNF-α, which activate NF-κB signaling. This heightened inflammation creates a pro-inflammatory environment in metabolic tissues, further disrupting normal metabolic processes and enhancing the risk of cardiovascular disease [78, 79].

Obesity driven by SIM1 dysfunction also fosters an inflammatory response in adipose tissue, where the accumulation of excess body fat encourages macrophage infiltration. These macrophages release additional cytokines that further activate NF-κB, creating a self-perpetuating feedback loop of inflammation. This cycle exacerbates metabolic dysregulation and perpetuates obesity, with long-term effects on systemic health, particularly in relation to cardiovascular and metabolic disorders [79, 80].

Furthermore, SIM1 dysfunction may contribute to increased oxidative stress due to the metabolic imbalances associated with obesity. Oxidative stress activates NF-κB through redox-sensitive pathways, intensifying systemic inflammation. This elevated oxidative stress, especially in the vascular endothelium, contributes to vascular damage and increases the risk of cardiovascular complications, reinforcing the connection between SIM1 variants, NF-κB activation, and cardiovascular disease [80, 81].

##### Downstream Effects of NF-κB Activation and Cardiovascular Disease (CVD)

Chronic NF-κB activation in individuals with SIM1-related obesity promotes the expression of adhesion molecules and chemokines in vascular tissues. These molecules attract immune cells to the endothelial surface, fostering the development of atherosclerotic plaques. The increased immune cell infiltration accelerates plaque formation, contributing to the narrowing of blood vessels and heightening the risk of cardiovascular events such as heart attacks and strokes [81, 82].

SIM1 variants are also linked to insulin resistance, a condition characterized by impaired cellular response to insulin. This insulin resistance is associated with increased NF-κB activation, which drives chronic inflammation. Elevated levels of circulating glucose and lipids further exacerbate this inflammatory state, worsening endothelial dysfunction and contributing to the progression of metabolic syndrome. The combined effects of insulin resistance and systemic inflammation promote cardiovascular disease by damaging the vascular endothelium and increasing the risk of atherosclerosis [82, 83].

Furthermore, NF-κB-mediated inflammation can impair endothelial function by reducing nitric oxide bioavailability, a key molecule responsible for blood vessel dilation and maintaining vascular health. Reduced nitric oxide availability leads to endothelial dysfunction, which increases vascular resistance and contributes to hypertension. This chronic elevation in blood pressure is a significant risk factor for cardiovascular complications, such as heart failure and stroke, thereby linking SIM1-related obesity, NF-κB activation, and cardiovascular disease [83, 84].

##### Implications for Cardiovascular Disease in the Context of Obesity

The connection between SIM1 variants, obesity, and NF-κB signaling highlights the central role of inflammation in the development of cardiovascular disease (CVD). Chronic NF-κB-driven inflammation exacerbates key pathological processes such as atherosclerosis, insulin resistance, and endothelial dysfunction, all of which contribute to the progression of CVD. The persistent activation of NF-κB in response to obesity-related factors intensifies the inflammatory environment, which accelerates vascular damage and increases the risk of cardiovascular complications [84, 85].

To mitigate the cardiovascular risks associated with SIM1 variants, effective therapeutic strategies should focus on managing obesity and reducing inflammation. Directly targeting NF-κB signaling through anti-inflammatory agents may help reduce systemic inflammation and prevent its harmful effects on the cardiovascular system. Additionally, lifestyle interventions aimed at improving metabolic regulation—such as exercise, dietary changes, and weight management—could provide long-term benefits by reducing adiposity and restoring normal metabolic function. Approaches that focus on restoring hypothalamic function, which is impaired by SIM1 variants, may also help to prevent or reduce the obesity-driven inflammation and metabolic dysregulation that underlie the increased cardiovascular risk. These combined strategies may offer comprehensive solutions for individuals affected by SIM1-related cardiovascular risk [85].

#### 9. TBC1D1 (TBC1 Domain Family Member 1) Gene

Genetic variations in the TBC1D1 (TBC1 Domain Family Member 1) gene, which encodes a protein involved in glucose and lipid metabolism, have been linked to obesity and associated metabolic disorders. TBC1D1 is critical for insulin signaling and the regulation of glucose uptake in adipose and muscle tissues. Variants in this gene can affect the NF-κB (nuclear factor kappa-light-chain-enhancer of activated B cells) signaling pathway, which plays a central role in inflammation and cardiovascular disease (CVD) [86].

##### TBC1D1 Variants and NF-κB Activation

TBC1D1 is a key regulator of insulin signaling, particularly in its role in mediating the translocation of glucose transporter type 4 (GLUT4) to the cell membrane, facilitating glucose uptake. Variants in the TBC1D1 gene can impair this process, leading to reduced glucose uptake and an increase in lipogenesis. This disruption in glucose metabolism is often associated with metabolic conditions such as obesity and insulin resistance. The metabolic imbalance induced by TBC1D1 variants creates an environment prone to inflammation, including the activation of NF-κB signaling pathways, which are key mediators of the inflammatory response [86, 87].

Insulin resistance, a condition commonly linked to TBC1D1 dysfunction, is associated with chronic low-grade inflammation. Elevated levels of pro-inflammatory cytokines such as TNF-α and IL-6 are frequently observed in individuals with insulin resistance, and these cytokines are potent activators of NF-κB signaling. As a result, TBC1D1 variants that disrupt normal glucose metabolism can lead to a vicious cycle of heightened NF-κB activation, further exacerbating inflammation and contributing to the progression of metabolic and cardiovascular diseases [87, 88].

##### Mechanisms of NF-κB Activation by TBC1D1 Variants

Reduced TBC1D1 activity due to genetic variants can disrupt insulin signaling, leading to metabolic disturbances such as elevated circulating levels of free fatty acids and pro-inflammatory cytokines. This metabolic dysregulation fosters a state of insulin resistance, which is characterized by enhanced NF-κB activation. In turn, this increased NF-κB activity drives the expression of inflammatory mediators, creating a pro-inflammatory environment within adipose tissue and other metabolic organs. The resultant inflammation further impairs insulin sensitivity, perpetuating the cycle of metabolic dysfunction [88, 89].

Variants in TBC1D1 may also contribute to adipose tissue dysfunction, where the accumulation of excess adiposity leads to immune cell infiltration, particularly macrophages. These immune cells release additional cytokines, further activating NF-κB signaling. This continuous inflammatory activation exacerbates adipose tissue dysfunction and promotes the development of obesity-related metabolic disorders, such as insulin resistance and dyslipidemia [89, 90].

Additionally, metabolic disturbances caused by TBC1D1 variants can increase oxidative stress, a key driver of NF- κB activation. Oxidative stress activates NF-κB through redox-sensitive signaling pathways, which contribute to systemic inflammation. This inflammation, in turn, plays a critical role in cardiovascular damage, further linking TBC1D1 variants, NF-κB activation, and increased cardiovascular risk [90, 91].

##### Downstream Effects of NF-κB Activation and Cardiovascular Disease (CVD)

Chronic activation of NF-κB due to TBC1D1-related obesity contributes to vascular inflammation, enhancing the expression of adhesion molecules and chemokines in endothelial cells. This promotes the recruitment of immune cells to the vascular endothelium, accelerating the formation of atherosclerotic plaques. The progression of atherosclerosis increases the risk of cardiovascular events such as heart attacks and strokes, as plaque buildup can obstruct blood flow and lead to vascular damage [91, 92].

NF-κB activation also impairs endothelial function by reducing nitric oxide production, a molecule critical for maintaining blood vessel dilation and regulating blood pressure. This reduction in nitric oxide availability leads to vasoconstriction, which increases vascular resistance and elevates blood pressure. In individuals with obesity and metabolic dysregulation, this effect heightens the risk of hypertension, a major contributor to cardiovascular disease [92, 93].

Furthermore, the interaction between TBC1D1 variants and NF-κB signaling exacerbates systemic inflammation, which further promotes insulin resistance. This vicious cycle of inflammation and insulin resistance is particularly detrimental as insulin resistance is a well-established risk factor for the development of cardiovascular disease. The combined effects of inflammation, endothelial dysfunction, and insulin resistance amplify the risk of CVD, making individuals with TBC1D1-related metabolic disturbances more susceptible to cardiovascular complications [93, 94].

##### Implications for Cardiovascular Disease in the Context of Obesity

The relationship between TBC1D1 variants, NF-κB signaling, and inflammation highlights the critical role of metabolic dysfunction in the development of cardiovascular disease. Chronic inflammation, driven by NF-κB activation, is a key contributor to atherosclerosis, endothelial dysfunction, and insulin resistance—all of which significantly increase the risk of cardiovascular events. The persistent inflammatory environment fostered by these mechanisms accelerates the progression of cardiovascular pathology, making individuals with TBC1D1- related obesity particularly vulnerable to cardiovascular complications [94, 95].

Given the central role of NF-κB signaling in this process, targeting the NF-κB pathway or improving TBC1D1 function may provide effective therapeutic strategies for those at risk of obesity-related cardiovascular disease. Interventions aimed at reducing NF-κB activation could mitigate the inflammatory response and potentially slow the progression of atherosclerosis and other related conditions. Additionally, lifestyle changes that enhance insulin sensitivity, such as regular physical activity and dietary modifications, can help reduce systemic inflammation and lower cardiovascular risk in individuals with TBC1D1 variants. These approaches may serve as valuable components of a broader therapeutic strategy to combat obesity-related cardiovascular disease [95].

#### 10. ADRB3 (Beta-3 Adrenergic Receptor) Gene

Genetic variations in the ADRB3 (Beta-3 Adrenergic Receptor) gene, which encodes a receptor primarily involved in the regulation of lipolysis and thermogenesis in adipose tissue, have been associated with obesity and metabolic disorders. These variations can influence the NF-κB (nuclear factor kappa-light-chain-enhancer of activated B cells) signaling pathway, which is crucial for mediating inflammation and cardiovascular disease (CVD) [96].

##### ADRB3 Variants and NF-κB Activation

The ADRB3 receptor plays a crucial role in regulating energy expenditure and lipid metabolism, particularly by promoting lipolysis in adipocytes. Genetic variations in the ADRB3 gene can result in altered receptor activity, which disrupts these metabolic processes and contributes to the development of obesity. Obesity itself is often characterized by chronic low-grade inflammation, a condition that is closely associated with the activation of NF- κB signaling [96, 97].

Variants in ADRB3 that lead to impaired lipolytic activity can exacerbate the accumulation of fat tissue, which in turn promotes the release of pro-inflammatory cytokines such as TNF-α and IL-6. These cytokines activate NF-κB signaling, initiating a cascade of inflammatory responses. The persistent inflammation further exacerbates metabolic dysregulation and contributes to the development of obesity-related complications, highlighting the critical interplay between ADRB3 variants, NF-κB activation, and inflammation [97, 98].

##### Mechanisms of NF-κB Activation by ADRB3 Variants

ADRB3 variants can influence the secretion of adipokines, which are signaling molecules produced by adipose tissue. These adipokines play a key role in regulating inflammation and metabolism. Variations in ADRB3 activity may result in a decreased secretion of anti-inflammatory adiponectin and an increase in pro-inflammatory cytokines such as TNF-α and IL-6. Elevated levels of these cytokines can activate the NF-κB pathway, initiating a cascade of inflammation. This chronic inflammatory state is often observed in obesity, where altered adipokine profiles exacerbate metabolic dysfunction and promote systemic inflammation [98, 99].

Furthermore, obesity and metabolic disturbances associated with ADRB3 variants can increase oxidative stress. Reactive oxygen species (ROS), which accumulate in this context, are potent activators of NF-κB signaling. ROS-induced activation of NF-κB exacerbates the inflammatory response and is a contributing factor to the elevated cardiovascular risk observed in individuals with ADRB3-related metabolic dysfunction [99, 100].

Dysregulation of adrenergic signaling due to ADRB3 variants can negatively impact endothelial function. Impaired endothelial responses increase vascular inflammation, promoting NF-κB activation in the vascular endothelium. This process contributes to the development of atherosclerosis and the progression of cardiovascular disease, further emphasizing the significance of ADRB3 variants in modulating NF-κB-driven inflammation and cardiovascular risk [100, 101].

##### Downstream Effects of NF-κB Activation and Cardiovascular Disease (CVD)

Chronic activation of NF-κB due to inflammation promotes the expression of adhesion molecules and chemokines in endothelial cells, facilitating the recruitment of immune cells to the vascular endothelium. This process contributes to the formation of atherosclerotic plaques, a key factor in the development of cardiovascular events such as heart attacks and strokes. The ongoing recruitment of immune cells and the accumulation of plaque heighten the risk of cardiovascular disease [101, 102].

In addition to promoting atherosclerosis, persistent NF-κB activation impairs nitric oxide production, which is essential for maintaining proper vascular tone. Nitric oxide is a vasodilator that helps regulate blood vessel dilation. Reduced nitric oxide availability, as a result of NF-κB activation, leads to vasoconstriction and increased vascular resistance, contributing to hypertension. Hypertension is a significant risk factor for various cardiovascular conditions, including heart failure and stroke [102, 103].

The combination of ADRB3 variants, obesity, and NF-κB signaling also exacerbates systemic inflammation, which can further promote insulin resistance. Insulin resistance is a hallmark of metabolic syndrome, a condition strongly linked to cardiovascular disease. Elevated inflammatory cytokines and dysregulated metabolic processes worsen endothelial dysfunction, insulin sensitivity, and lipid profiles, further elevating the risk of cardiovascular complications in individuals with obesity and metabolic disturbances [103, 104].

##### Implications for Cardiovascular Disease in the Context of Obesity

The connection between ADRB3 variants, NF-κB activation, and inflammation underscores the significant role of metabolic dysfunction in the development of cardiovascular disease (CVD). Chronic inflammation driven by NF-κB activation plays a central role in the progression of atherosclerosis, endothelial dysfunction, and insulin resistance—key factors that increase the risk of CVD. The elevated levels of pro-inflammatory cytokines and disrupted metabolic processes observed in obesity further exacerbate vascular damage, promoting plaque formation and contributing to hypertension, insulin resistance, and other metabolic disturbances that heighten cardiovascular risk [104, 105].

Therapeutic strategies targeting the ADRB3 pathway or NF-κB signaling could provide promising interventions for individuals at risk of obesity-related cardiovascular disease. By enhancing adrenergic signaling through ADRB3 agonists, promoting lipolysis, and reducing excess adiposity, it may be possible to improve metabolic health and decrease the inflammatory burden. Additionally, directly targeting the NF-κB pathway with anti-inflammatory agents or other therapies could reduce systemic inflammation and its detrimental effects on cardiovascular and metabolic health. Lifestyle interventions, such as physical activity and dietary changes, may also play a crucial role in improving both adrenergic signaling and inflammatory responses, thereby reducing cardiovascular risks associated with obesity [105].

#### 11. UCP1 (Uncoupling Protein 1) Gene

Genetic variations in the UCP1 (Uncoupling Protein 1) gene, which encodes a mitochondrial protein primarily involved in thermogenesis and energy expenditure in brown adipose tissue, have significant implications for metabolism, obesity, and cardiovascular health. UCP1 plays a critical role in dissipating energy as heat and regulating metabolic processes. Variations in the UCP1 gene can influence the NF-κB (nuclear factor kappa-light-chain-enhancer of activated B cells) signaling pathway, which is central to inflammation and cardiovascular disease (CVD) [106].

##### UCP1 Variants and NF-κB Activation

The uncoupling protein 1 (UCP1) plays a critical role in maintaining energy homeostasis by facilitating adaptive thermogenesis. This process is particularly important during cold exposure and periods of overfeeding, where UCP1 enables the dissipation of excess energy as heat. Genetic variations in UCP1 can influence its expression and functional capacity, potentially altering energy expenditure and fat storage dynamics. A diminished thermogenic response may predispose individuals to obesity, a condition often linked to chronic low-grade inflammation and increased activation of nuclear factor kappa B (NF-κB) [106, 107].

Obesity is closely associated with an inflammatory state characterized by the upregulation of NF-κB signaling pathways. Impaired UCP1-mediated thermogenesis, as a result of genetic variants, may exacerbate adiposity. The resulting expansion of adipose tissue promotes the release of pro-inflammatory cytokines, which in turn activate NF-κB. This heightened NF-κB activity contributes to systemic inflammation, establishing a potential mechanistic link between UCP1 variants, thermogenic dysfunction, and inflammation observed in obesity [107, 108].

##### Mechanisms of NF-κB Activation by UCP1 Variants

UCP1 plays a critical role in maintaining adipose tissue homeostasis. Genetic variants that reduce UCP1 function may contribute to the dysfunction of brown adipose tissue, characterized by lipid accumulation and the release of inflammatory mediators. This dysfunction triggers the activation of NF-κB signaling, driven by cytokines such as tumor necrosis factor-alpha (TNF-α) and interleukin-6 (IL-6). These cytokines perpetuate the inflammatory response, establishing a cycle that reinforces tissue inflammation and systemic immune activation [108, 109].

Variations in UCP1 also impact mitochondrial function, particularly in the generation and regulation of reactive oxygen species (ROS). Impaired UCP1 activity may lead to elevated oxidative stress, a known activator of NF-κB. Increased ROS levels promote the activation of NF-κB signaling, exacerbating inflammation and contributing to the pathophysiology of cardiovascular diseases. This oxidative stress-induced inflammation underscores the critical role of mitochondrial integrity in metabolic and cardiovascular health [109, 110].

In addition to its role in thermogenesis, UCP1 is integral to energy expenditure and metabolic regulation. Genetic variants that impair thermogenic capacity can result in energy imbalance and increased adiposity. The resultant metabolic changes intensify inflammatory signaling through NF-κB activation, creating a feedback loop of inflammation. This chronic inflammatory state not only worsens obesity but also heightens cardiovascular risk, highlighting the broader implications of UCP1 variants in systemic health [110, 111].

##### Downstream Effects of NF-κB Activation and Cardiovascular Disease (CVD)

Chronic activation of NF-κB in the context of obesity has significant implications for vascular health. One critical outcome is the promotion of atherosclerosis through enhanced expression of adhesion molecules and pro-inflammatory cytokines in vascular tissues. These molecules facilitate the recruitment and retention of immune cells at the endothelium, initiating and propagating the formation of atherosclerotic plaques. Over time, this inflammatory milieu increases the risk of cardiovascular events, including myocardial infarction and stroke [111,112].

NF-κB activation also impairs endothelial function by disrupting nitric oxide (NO) bioavailability. NO is a key regulator of vascular tone, and its depletion leads to vasoconstriction and elevated blood pressure. Persistent hypertension, driven by reduced NO and chronic NF-κB activity, is a major contributor to cardiovascular disease, exacerbating the burden of vascular complications associated with obesity [112, 113].

The systemic effects of NF-κB activation extend to metabolic processes, where inflammation-induced insulin resistance emerges as a critical factor. The interplay between UCP1 variants, heightened NF-κB signaling, and obesity amplifies systemic inflammation, further impairing insulin sensitivity. This cascade increases the risk of metabolic syndrome, a cluster of conditions that significantly elevates the likelihood of cardiovascular disease. The interconnected nature of these pathways underscores the importance of addressing chronic inflammation and metabolic dysfunction in reducing cardiovascular risk [113, 114].

##### Implications for Cardiovascular Disease in the Context of Obesity

The interplay between UCP1 variants, NF-κB signaling, and inflammation underscores the critical role of metabolic dysfunction in the development of cardiovascular disease (CVD). Chronic inflammation mediated by persistent NF-κB activation drives key pathological processes, including endothelial dysfunction, insulin resistance, and atherosclerosis. These mechanisms collectively heighten cardiovascular risk, particularly in individuals with obesity, where impaired thermogenesis and increased adiposity exacerbate the inflammatory state [114, 115].

Addressing these pathways presents potential therapeutic opportunities for mitigating obesity-related cardiovascular disease. Enhancing UCP1 function through pharmacological or lifestyle interventions could improve thermogenesis and energy balance, reducing adiposity and systemic inflammation. Targeting NF-κB signaling directly offers another avenue to curb chronic inflammation and its downstream effects on vascular and metabolic health. Strategies aimed at promoting weight loss, improving endothelial function, and attenuating inflammatory signaling may collectively reduce cardiovascular risk in this vulnerable population, offering a multidimensional approach to managing obesity-related CVD [115].

#### 12. SH2B1 (SH2B Adaptor Protein 1) Gene

Genetic variations in the SH2B1 (SH2B Adaptor Protein 1) gene have been implicated in obesity, insulin resistance, and related metabolic disorders. SH2B1 is an adaptor protein that plays a key role in the signaling pathways of insulin and leptin, which are critical for energy homeostasis. Additionally, SH2B1 has been linked to the regulation of the NF-κB (nuclear factor kappa-light-chain-enhancer of activated B cells) signaling pathway, which is essential for mediating inflammation and cardiovascular disease (CVD) [116].

##### SH2B1 Variants and NF-κB Activation

The SH2B1 protein plays a critical role in amplifying insulin and leptin signaling by facilitating the activation of key pathways, including phosphoinositide 3-kinase (PI3K) and Akt. These pathways are essential for maintaining metabolic homeostasis and energy balance. Genetic variants in the SH2B1 gene can impair its function, leading to diminished sensitivity to insulin and leptin. This dysregulation disrupts metabolic signaling and predisposes individuals to obesity, a condition characterized by chronic low-grade inflammation and heightened activation of nuclear factor kappa B (NF-κB) [116, 117].

The metabolic dysregulation associated with SH2B1 variants contributes to the accumulation of adipose tissue, which serves as a significant source of pro-inflammatory cytokines such as tumor necrosis factor-alpha (TNF-α) and interleukin-6 (IL-6). These cytokines activate the NF-κB pathway, amplifying the inflammatory response.

Genetic variations in SH2B1 that exacerbate metabolic imbalances further enhance cytokine production, creating a feedback loop of inflammation and NF-κB activation. This pathway not only reinforces the chronic inflammatory state associated with obesity but also contributes to the development of related metabolic and cardiovascular complications [117, 118].

##### Mechanisms of NF-κB Activation by SH2B1 Variants

Genetic variations in SH2B1 disrupt the signaling cascades initiated by insulin and leptin, two critical regulators of energy and metabolic homeostasis. Impaired SH2B1 function compromises these pathways, leading to increased lipogenesis and reduced lipolysis, which drive the development of obesity. The excessive adiposity promotes the secretion of inflammatory mediators, triggering the activation of the nuclear factor kappa B (NF-κB) signaling pathway. This activation perpetuates a cycle of inflammation that exacerbates metabolic dysfunction [118, 119].

Adipocyte dysfunction is another consequence of SH2B1 variants, contributing to chronic inflammation. Dysfunctional adipocytes attract immune cells, particularly macrophages, to adipose tissue. These recruited immune cells release pro-inflammatory cytokines such as TNF-α and IL-6, which activate NF-κB and sustain the inflammatory state. This creates a feedback loop where inflammation further promotes immune cell recruitment and cytokine release, compounding the dysregulation of adipose tissue [119, 120].

In addition to inflammation, oxidative stress is a key mechanism linking SH2B1 variants to NF-κB activation. Metabolic disturbances caused by impaired SH2B1 function lead to excessive production of reactive oxygen species (ROS). Oxidative stress serves as a potent activator of NF-κB, amplifying inflammatory signaling and reinforcing the pro-inflammatory environment. Together, these mechanisms highlight the multidimensional role of SH2B1 variants in promoting chronic inflammation and its downstream metabolic and cardiovascular effects [120, 121].

##### Downstream Effects of NF-κB Activation and Cardiovascular Disease (CVD)

Chronic activation of nuclear factor kappa B (NF-κB) in response to inflammation has significant implications for vascular health, particularly through its role in endothelial dysfunction. NF-κB promotes the upregulation of adhesion molecules on endothelial cells, facilitating the recruitment of immune cells to vascular walls.

Simultaneously, it reduces nitric oxide (NO) bioavailability, impairing the vasodilatory function of the endothelium. This dysfunction is a critical precursor to atherosclerosis and other cardiovascular conditions [121,122].

NF-κB signaling plays a central role in the development and progression of atherosclerosis. Inflammatory cytokines produced by NF-κB activation attract monocytes to the endothelial layer, where they differentiate into macrophages and engulf oxidized lipids, forming foam cells. The accumulation of foam cells contributes to the formation of atherosclerotic plaques, further exacerbating vascular inflammation and narrowing arterial walls, which increases the risk of cardiovascular events [122, 123].

Hypertension is another consequence of NF-κB-mediated inflammation. The chronic inflammatory response can induce vascular remodeling, characterized by structural changes in blood vessels that increase stiffness and resistance. This process, coupled with impaired NO-mediated vasodilation, elevates blood pressure.

Hypertension is particularly prevalent in individuals with obesity and metabolic syndrome, linking NF-κB activation to the broader spectrum of cardiovascular complications associated with metabolic dysfunction [123,

124].

##### Implications for Cardiovascular Disease in the Context of Obesity

The interaction between SH2B1 variants, NF-κB signaling, and chronic inflammation underscores the central role of metabolic dysfunction in the pathogenesis of cardiovascular disease (CVD). Variants in SH2B1 impair insulin and leptin signaling, leading to obesity and a cascade of metabolic disturbances. These disturbances drive persistent low-grade inflammation through NF-κB activation, which promotes the development of atherosclerosis, insulin resistance, and other cardiovascular risks associated with obesity. The inflammatory state exacerbates vascular dysfunction and metabolic instability, creating a compounding effect that heightens the likelihood of cardiovascular complications [124, 125].

Therapeutic strategies targeting the SH2B1 pathway or NF-κB signaling hold promise for reducing obesity-related cardiovascular risk. Enhancing insulin and leptin sensitivity through pharmacological or lifestyle interventions could restore metabolic balance and reduce adiposity. Additionally, therapies aimed at attenuating NF-κB activity may help to mitigate inflammation and its downstream vascular effects. Addressing both metabolic dysregulation and inflammation through such targeted approaches could provide a comprehensive strategy for reducing cardiovascular disease burden in individuals with obesity [125].

## Discussion

The complex relationship between genetic variations in obesity-related genes and the NF-κB signaling pathway reveals significant implications for understanding the pathophysiology of obesity and its associated cardiovascular diseases. Our investigation illustrates how genetic factors not only predispose individuals to obesity but also perpetuate a cycle of inflammation that contributes to cardiovascular pathology. By elucidating the mechanisms through which these genetic variations influence NF-κB activation, we can better understand the underlying processes that exacerbate inflammation and metabolic dysregulation in obese individuals.

The findings highlight that specific genes, such as FTO and MC4R, are crucial in modulating energy homeostasis and appetite regulation. Variations in these genes can lead to altered signaling pathways that enhance NF-κB activation, resulting in increased secretion of pro-inflammatory cytokines. This inflammatory state is further aggravated by genetic variations in LEP and LEPR, which disrupt leptin signaling and adipocyte function. Such dysregulation fosters an environment conducive to chronic inflammation, a hallmark of obesity that predisposes individuals to cardiovascular disease.

Additionally, the role of other genes, such as SH2B1 and UCP1, underscores the complexity of metabolic pathways involved in obesity and inflammation. SH2B1 variants can impair insulin and leptin signaling, leading to increased adiposity and enhanced NF-κB activation. Meanwhile, UCP1 variations can disrupt thermogenic function, contributing to oxidative stress and further NF-κB activation. These interactions collectively emphasize the necessity of a multifactorial approach to understanding obesity, one that considers the interplay between genetic predispositions and environmental factors.

The implications of our findings extend beyond mere understanding; they inform potential therapeutic strategies. Targeting the NF-κB pathway could offer new avenues for intervention in the prevention and treatment of obesity-related cardiovascular disease. Developing pharmacological agents that inhibit NF-κB activation or enhance insulin and leptin sensitivity may mitigate inflammation and improve metabolic outcomes. Additionally, lifestyle interventions aimed at reducing adiposity and enhancing energy expenditure may also serve to decrease NF-κB-mediated inflammation, thereby lowering cardiovascular risk.

Moreover, this investigation underscores the importance of personalized medicine in obesity treatment. Understanding an individual’s genetic makeup could guide tailored interventions, optimizing therapeutic efficacy and improving health outcomes. By integrating genetic profiling into clinical practice, healthcare providers could better predict obesity-related complications and develop customized prevention strategies.

Our investigation emphasizes the critical role of genetic variations in obesity-related genes in influencing NF-κB signaling and its downstream inflammatory effects. The interplay between these genetic factors and the inflammatory response presents significant implications for cardiovascular health in obese individuals. Continued research is essential to further elucidate these relationships and explore innovative therapeutic approaches that address the complex challenges posed by obesity and its comorbidities.

## Key findings

This study explores the relationship between genetic variations in obesity-related genes and the activation of the NF-κB signaling pathway, with a focus on cardiovascular disease risk. Specific genetic variants in genes such as FTO, MC4R, LEP, LEPR, and SH2B1 were found to be associated with heightened NF-κB activation. These genetic changes contribute to the disruption of insulin and leptin signaling pathways, both of which are crucial for maintaining energy balance and regulating inflammatory processes.

The investigation also revealed that these genetic variations increase the production of pro-inflammatory cytokines, which in turn sustain NF-κB activation. This chronic inflammatory state creates a harmful feedback loop, exacerbating metabolic dysfunction and increasing the risk of cardiovascular disease. Additionally, genetic variants in PPARG and UCP1 were identified as significant contributors to oxidative stress, a factor that further intensifies NF-κB signaling. This highlights the role of metabolic dysregulation in promoting inflammation within adipose tissue.

Persistent NF-κB activation, driven by genetic predispositions, leads to endothelial dysfunction, a key early event in the development of atherosclerosis. These findings underline the crucial role of obesity-related genetic factors in the onset of cardiovascular complications. The study’s results also point to the potential for personalized medicine in addressing obesity-related inflammation and cardiovascular risk. By considering individual genetic profiles, therapeutic strategies could be tailored to target NF-κB signaling pathways, offering more effective interventions for populations at risk.

## Conclusion

This comprehensive investigation into the genetic variations of obesity-related genes—specifically FTO, MC4R, LEP, LEPR, PCSK1, PPARG, BDNF, SIM1, TBC1D1, ADRB3, UCP1, and SH2B1—has illuminated their critical roles in modulating the NF-κB signaling pathway. The alterations in these genes contribute to dysregulation of key metabolic processes, leading to an increased propensity for obesity and its associated complications.

It was observed that genetic variations in these genes can impact NF-κB activation through various mechanisms, including impaired insulin and leptin signaling, altered adipocyte function, increased oxidative stress, and heightened secretion of pro-inflammatory cytokines. The chronic activation of the NF-κB pathway as a result of these variations drives systemic inflammation, which is a hallmark of obesity-related pathologies.

The downstream effects of NF-κB activation manifest in several detrimental ways, particularly in the context of cardiovascular disease. This includes endothelial dysfunction, promotion of atherosclerosis, and the exacerbation of hypertension, all of which are aggravated by chronic inflammation associated with obesity. The delicate balance between genetic predispositions and inflammatory responses underscores the complexity of obesity as a multifactorial disease, with significant implications for cardiovascular health.

Given the significant role of NF-κB signaling in mediating inflammation and cardiovascular pathology, understanding these genetic interactions opens avenues for targeted therapeutic strategies. Interventions aimed at mitigating NF-κB activation, improving metabolic function, and reducing inflammation may provide effective means to address the increased cardiovascular risk associated with obesity. This emphasizes the need for further research into the genetic underpinnings of obesity and their contribution to cardiovascular disease, providing insights that could inform personalized approaches to prevention and treatment.

## Abbreviations

FTO: Fat Mass and Obesity-Associated Gene
MC4R: Melanocortin-4 Receptor Gene
LEP: Leptin Gene
LEPR: Leptin Receptor Gene
PCSK1: Proprotein Convertase Subtilisin/Kexin Type 1 Gene
PPARG: Peroxisome Proliferator-Activated Receptor Gamma Gene
BDNF: Brain-Derived Neurotrophic Factor Gene
SIM1: Single-Minded Homolog 1 Gene
TBC1D1: TBC1 Domain Family Member 1 Gene
ADRB3: Beta-3 Adrenergic Receptor Gene
UCP1: Uncoupling Protein 1 Gene
SH2B1: SH2B Adaptor Protein 1 Gene
NF-κB: Nuclear Factor Kappa-Light-Chain-Enhancer of Activated B Cells
CVD: Cardiovascular Disease

## Declarations

### Ethics declarations

Ethics approval and consent to participate

Not applicable.

## Consent for publication

Not applicable.

## Data Availability statement

All data generated or analyzed during this study are included in this article.

## Funding

I declare that there was not any source of funding for this research work.

### Acknowledgements

“Not applicable”.

## Authors’ Information

1. Viqas Shafi (VS) is the author of the study and contributed to its conceptualization, design, and methodology, as well as the literature search and referencing. He was responsible for writing, editing, and revising the manuscript, as well as delineating the findings, results, conclusions, implications, and all other aspects of the study. VS conducted data extraction and analysis, critically evaluated every aspect of the study, ensured adherence to relevant PRISMA guidelines, and addressed study limitations and references. Additionally, he created PRISMA Flow Diagram. The author reviewed and approved the manuscript. He investigated how genetic variations in obesity-related genes, including FTO, MC4R, LEP, LEPR, PCSK1, PPARG, BDNF, SIM1, TBC1D1, ADRB3, UCP1, and SH2B1, impact the NF-κB signaling pathway, focusing on mechanisms of NF-κB activation, downstream inflammatory responses, and their contribution to cardiovascular disease development in the context of obesity. Viqas Shafi (VS), Doctor of Pharmacy (Pharm.D.) from Dow University of Health Sciences, Karachi, Pakistan, focuses his research on disease mechanisms, pharmacology, pharmacotherapy, pharmacogenetics, precision medicine, molecular and cellular biology, and cell signaling pathways.
2. Nabeel Ahmad Khan (NAK) is the co-author of the study and contributed to the writing, editing and revision. He contributed to the results, discussion and conclusion sections of the study along with working on the findings, interpretation of the data and references. He contributed to investigating the impact of genetic variations in obesity-related genes (FTO, MC4R, LEP, LEPR, and PCSK1) on the NF-κB signaling pathway, exploring mechanisms of NF-κB activation, downstream inflammatory responses, and their role in cardiovascular disease development in the context of obesity. Nabeel Ahmad Khan (NAK) holds a Doctor of Pharmacy (Pharm.D.) degree from Dow University of Health Sciences, Karachi, Pakistan, completed in 2015, and a Master’s in Multidisciplinary Biomedical Sciences with a concentration in pharmacology from the University of Alabama at Birmingham, completed in 2022. His research interests include exploring the causal link between environmental factors and disease pathogenesis, disease mechanisms and cell signaling pathways. Through his work, he seeks to advance the field of biomedical sciences by uncovering critical insights that can lead to improved disease prevention and treatment.
3. Ifrah Siddiqui (IS)* is the co-author of the study and contributed to the writing, editing and revision. She contributed to the results, discussion and conclusion sections of the study along with working on the findings, interpretation of the data and references. She contributed to investigating the impact of genetic variations in obesity-related genes (FTO, MC4R, LEP, LEPR, PCSK1, PPARG, BDNF, SIM1, TBC1D1, ADRB3, UCP1, and SH2B1) on the NF-κB signaling pathway, exploring mechanisms of NF-κB activation, downstream inflammatory responses, and their role in cardiovascular disease development in the context of obesity. Ifrah Siddiqui (IS)* holds a Bachelor’s Degree with a focus on Psychology from the University of Karachi, Pakistan. Currently, she is undertaking training/course in Health & Medicine from Harvard Medical School. She has a passion for investigating the molecular mechanisms underlying disease pathogenesis and psychological aspects of various diseases.

Corresponding author: IS

Correspondence to Ifrah Siddiqui

The work and contributions of everyone have been described in detail, the order is randomized and the numbering is just for referencing purpose.

